# Mediating Role of Depression and Anxiety in the Association Between Food Insecurity and Delayed TB Treatment in Botswana: A Cross-Sectional Study

**DOI:** 10.64898/2026.04.08.26350465

**Authors:** Emmanuel Sakyi, Keneilwe Molebatsi, Chawangwa Modongo, Sanghyuk S. Shin

## Abstract

**Background:** Delayed TB treatment remains a major challenge to tuberculosis (TB) control in low- and middle-income countries, where it contributes to prolonged infection, higher costs, and increased drug resistance. Food insecurity is an important determinant of health among people with TB and may contribute to delayed treatment through economic, physical, and psychosocial pathways. Depression and anxiety have also been associated with both food insecurity and delayed TB treatment, suggesting that they may mediate this relationship. This study examined the association between food insecurity and delayed TB treatment initiation and assessed the mediating roles of depression for this relationship among people newly diagnosed with TB in Botswana.

**Methods:** We recruited 180 participants newly diagnosed with TB in Gaborone, Botswana. Food insecurity, depression, and anxiety were measured using the Household Food Insecurity Access Scale, PHQ-9, and Zung Self-Rating Anxiety Scale, respectively. Logistic regression was used to examine the association between food insecurity and delayed TB treatment. Mediation analysis using a counterfactual framework was conducted to assess the mediating roles of depression and anxiety.

**Results:** Among the 180 participants, 45 (25%) experienced delayed TB treatment initiation. Participants with delayed TB treatment had higher mean scores for food insecurity (4.16 vs 2.99, p = 0.11), depression (10.1 vs 6.92, p = 0.001), and anxiety (37.1 vs. 33.9, p = 0.05). While standard regression analysis found insufficient evidence of an overall association between food insecurity and delayed TB treatment initiation (OR = 1.04, 95% CI: 0.98–1.11, p = 0.20), mediation analysis found evidence of total natural indirect effects through depression (OR = 1.04, 95% CI: 1.01-1.08, p<0.001, 99% mediated) and anxiety (OR = 1.03, 95% CI: 1.01 - 1.06, p<0.001, 38% mediated).

**Conclusion:** Mediation analysis revealed that depression and anxiety may represent pathways linking food insecurity to delayed TB treatment. These findings highlight the importance of considering both socioeconomic and psychological factors in addressing delayed TB treatment. Further studies are needed to confirm these pathways.

## Background

Delayed TB treatment poses a major barrier to tuberculosis (TB) control (1). Delayed treatment often leads to severe TB symptoms, prolonged duration of TB infection, increased cost of treatment, and increased mortality, especially among people living with HIV (2,3). Delayed TB treatment has also been reported as a contributing factor for development of resistance to anti-tuberculosis drugs which hinders public health efforts to end TB (4–6). Understanding specific factors associated with delayed TB treatment is therefore important to guide strategies to improve treatment initiation and control programs.

Prior studies have reported depression and anxiety as important factors associated with delayed TB treatment and treatment non-adherence leading to poor outcomes (7,8). Depression and anxiety present with symptoms such as hopelessness, excessive fear, worry, low self-worth, lack of motivation, and loss of interest in activities. These symptoms may reduce the motivation and drive to seek and remain engaged in TB care leading to delay in TB diagnosis and treatment (9).

Food insecurity is recognized as an important determinant of health among people with TB and or HIV (10). Food insecurity refers to the condition in which the people do not have a stable access to safe and sufficient quantity of food that meets their nutritional needs (11,12). Food insecurity may contribute to delayed TB diagnosis and treatment through several economic and physical pathways. Individuals experiencing food insecurity may lack the financial resources needed for transport to health facilities, consultation fees and other indirect care seeking costs (10,13,14). This challenge may be worsened by coexisting conditions such as unemployment, low-income jobs and residence in poorer communities far from health care sites (15). For example, in the study by Teo and colleagues, participants often cited distance from health facilities and financial constraints as the reasons for delay in seeking TB treatment (13). Subsequent poor nutritional status from food insecurity worsens treatment outcomes especially for those with HIV coinfection (16,17). Recent studies have also reported strong association between food insecurity and mental health outcomes particularly depression and anxiety (18,19). Soboka et al. reported four times higher odds of mental distress among those experiencing food insecurity in southwest Ethiopia (18). These findings suggest that depression and anxiety may act as potential pathways through which food insecurity influences delayed TB treatment.

Despite these established relationships, the potential pathways mediating the association between food insecurity and delayed TB treatment have not been well examined. Understanding this association and its mediated pathways is important for public health, as food insecurity is a modifiable social determinant and understanding the physical, economic and psychosocial pathways could inform the development of interventions aimed at reducing delays in TB treatment initiation and improving TB outcomes. Therefore, this study sought to examine the association between food insecurity and delayed TB treatment and the mediating role of depression or anxiety for this relationship among people newly diagnosed with TB in Botswana.

## Methods

### Study design and population

This study analyzed data from a 2019 cross-sectional research project conducted across 12 health centers in Gaborone, Botswana. Details of the study procedures can be found in previous studies from this survey (8,19). This current study examines the association between food insecurity and delayed TB treatment initiation and whether depression or anxiety mediates this relationship.

### Study population

Research assistants approached people who were 18 years old or more, newly diagnosed with TB and receiving care across the 12 health centers. Potential participants were assessed for eligibility, and TB diagnosis was confirmed by reviewing history from clinic records. Face-to-face interviews were used to obtain written-informed consent after assessing eligibility. Questionnaires were then administered by research assistants in local Setswana language or English based on participant preference. Ethical approval for the study was obtained from the Institutional Review Boards at the Botswana Ministry of Health and Wellness and University of California Irvine. This study was conducted according to the guidelines of the Helsinki Declarations.

### Measures

Sociodemographic data such as participant sex, age, educational status, and marital status were collected using researcher designed questionnaires. Other clinical information such as TB classification, TB symptoms, duration of symptoms, HIV status, CD-4 T cell count, and antiretroviral history were also collected from clinic records. Food insecurity, depression and anxiety were measured using standardized measurement tools.

In the current study, delayed TB treatment was defined as experiencing TB symptoms for 2 months or more before TB treatment initiation. Participants who reported any TB symptom such as fever, hemoptysis, weight loss, cough, and night sweat for a 2-month duration or more was classified as delayed TB treatment initiation.

Food insecurity was measured using the Household Food Insecurity Access Scale (HFIAS) developed by the USAID (20). The nine-item HFIAS questionnaire has been widely validated for assessing the degree of food insecurity within households over the previous four weeks (20). The HFIAS achieved a Cronbach’s alpha of 0.94 within our study sample. Participant responses were recorded with a scale ranging from Never (0) to often (3); for total scale interpretation, higher scores indicate higher food insecurity with possible range of 0 to 27.

We measured depression among participants using the nine-item Patient Health Questionnaire-9 (PHQ-9). The PHQ-9 is also validated for screening depressive symptoms in many settings including Botswana (8,21). The total scores on the PHQ-9 range from 0 to 27 with higher sores indicating higher depressive symptoms. The internal reliability score for the PHQ-9 was acceptable among our study sample (Cronbach’s alpha=0.74).

Lastly, the twenty-item ZUNG Self-Rating Anxiety Scale (ZUNG) was used to screen for anxiety symptoms. The ZUNG allows participants to select how much an item applies to them on a scale of 1 to 4 (22–24). The Zung scores range from 20 to 80; higher scores indicate more severe anxiety symptoms. The ZUNG achieved a Cronbach’s alpha of 0.80 within our study sample.

## Statistical analysis

R Statistical Software version 4.5.1 was used to perform all data analysis. We used descriptive tables and figures to summarize the continuous variables using means and medians whereas frequencies and percentages were used for categorical variables. We used the two-sample t-test to compare the distribution of normally distributed continuous variables between delayed and not delayed TB treatment initiation categories and the Wilcoxon Rank Sum test for continuous variables that were not normally distributed.

We first examined the association between food insecurity and delayed TB treatment using standard logistic regression modelling. Food insecurity was analyzed as a continuous variable using the sum scores on the HFIAS whereas delayed TB treatment initiation was modelled as a binary categorical variable (delayed or not delayed). We fitted bivariate logistic regression models between exposure and outcome variable to test for independent association. We used a conceptual diagram of causal determinants to identify possible confounders (age, sex, marital status, income status, educational status, HIV status) (**Figure 1**, which shows a conceptual model for our mediation analysis). Participants who were widowed or divorced were classified as single for the analysis.

**Figure 1.**
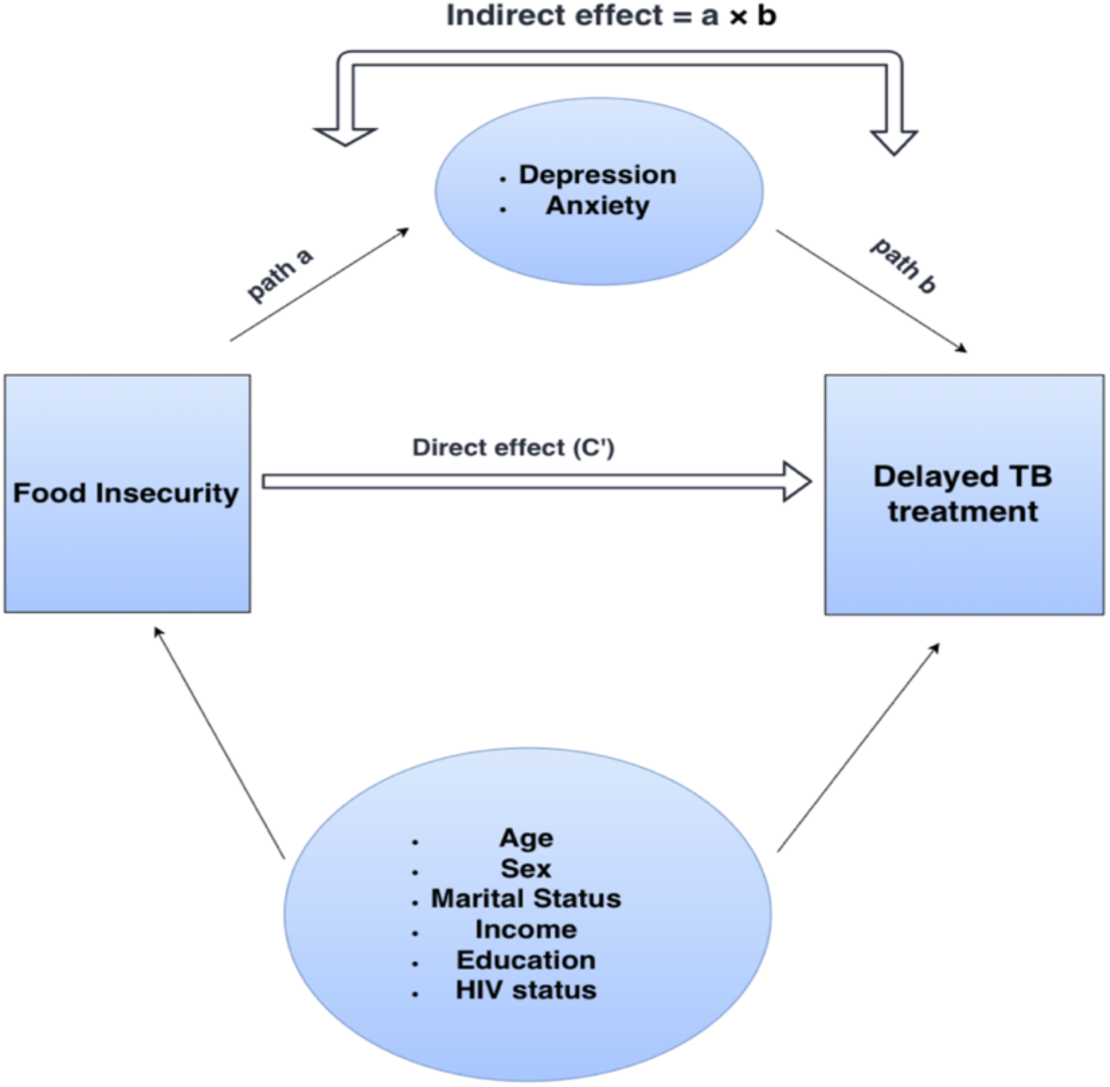
Conceptual Model

Due to the limited number of delayed TB treatment events in our data, we selected variables to include in the final model using the change-in-estimate method (25). Independent bivariate models were fitted for each possible confounder and the outcome variable. Variables were added in a stepwise manner to the initial bivariate model for food insecurity and delayed TB treatment. Variables were added starting from those with the lowest p-value for the bivariate associations with the outcome variable. Using the change-in-estimate method, a variable that caused a change of 10% or more in odds ratio (OR) for food insecurity was retained as a confounder for the final model. After applying the change-in-estimate method, none of the variables caused a 10% change in OR hence they were all dropped. Therefore, the initial bivariate model for food insecurity and delayed TB treatment initiation was maintained as the final model. A p-value < 0.05 was considered statistically significant. We conducted sensitivity analysis by including HIV status in the final model regardless of its confounding effect.

Next, we tested whether depression or anxiety lies on the pathway between food insecurity and delayed TB treatment initiation using mediation analysis based on a counterfactual framework to decompose the overall association into direct and indirect effects (26). Mediation analysis was done regardless of the significance of the total effect according to current recommendations for mediation analysis (27,28). Separate mediation analysis was done for depression and anxiety using the Cmest function in the CMAverse R package (29). Exposure– mediator interaction was assessed by including an interaction term between the exposure and each mediator. Evidence of interaction was observed for anxiety but not depression, hence interaction was specified for the anxiety mediation model. Pure natural direct effect is the direct effect of food insecurity on delayed TB given the mediation is fixed at the control level. Total natural indirect effect is the effect of food insecurity on delayed TB when the mediation variable is allowed to change due to food insecurity. Total effect is the sum of pure natural direct effect and the total natural indirect effect (30,31). Bootstrap method was used to estimate the 95% confidence intervals for causal mediation effects. The mediation analyses were reported in accordance with “A Guideline for reporting Mediation Analyses (AGReMA) statement” (**additional file 1**) (32). Mediation analysis in this study assumed no unmeasured confounding strong enough to affect the results. We also assumed a temporal ordering, where household food insecurity preceded mental health outcomes, and mental health outcomes preceded delayed TB treatment.

## Results

The overall characteristics of study participants is shown in **Table 1**. The study recruited 180 people newly diagnosed with TB, with most participants being males (64.4%) and single (90%). Among the 180 participants, 45 (25%) experienced delayed TB treatment initiation. The mean age of participants was 38.2 years with those who had delayed TB treatment initiation being older (40.4 years vs. 37.5 years). A greater percentage of the study participants (60.6%) had less than secondary school education and earned some form of income (63.9%). Over half (55.0%) of study participants tested positive for HIV. Among those that tested positive for HIV, the mean CD4 T cell count was 341. The mean food insecurity scores were higher among those who experienced delayed TB treatment (4.16 vs. 2.99, p = 0.11), (**Figure 2**, which shows the distribution of food insecurity scores by TB treatment initiation). Participants who experienced delay in TB treatment also reported higher mean depression scores compared to those who did not report delayed treatment (10.10 vs 6.92, p = 0.001) (**Figure 3**, which shows the distribution of depression scores by TB treatment initiation). Similarly, the mean anxiety scores were also higher among those with delayed TB treatment initiation (37.10 vs. 33.90, p = 0.05) (**Figure 4**, which shows the distribution of anxiety scores by TB treatment initiation).

**Table 1:** Characteristics of Study Participants by TB Treatment Initiation.

|  | Not delayed<br>(N=135) | delayed<br>(N=45) | Overall<br>(N=180) |
| --- | --- | --- | --- |
| **Sex** |  |  |  |
| Female | 48 (35.6%) | 16 (35.6%) | 64 (35.6%) |
| Male | 87 (64.4%) | 29 (64.4%) | 116 (64.4%) |
| **Age** |  |  |  |
| Mean (SD) | 37.5 (12.2) | 40.4 (13.8) | 38.2 (12.6) |
| **Marital Status** |  |  |  |
| Single | 122 (90.4%) | 40 (88.9%) | 162 (90.0%) |
| Married | 13 (9.6%) | 5 (11.1%) | 18 (10.0%) |
| **Educational status** |  |  |  |
| Less than secondary | 81 (60.0%) | 28 (62.2%) | 109 (60.6%) |
| Secondary or above | 54 (40.0%) | 17 (37.8%) | 71 (39.4%) |
| **Income status** |  |  |  |
| None | 49 (36.3%) | 16 (35.6%) | 65 (36.1%) |
| Yes | 86 (63.7%) | 29 (64.4%) | 115 (63.9%) |
| **HIV Status** |  |  |  |
| Negative | 64 (47.4%) | 17 (37.8%) | 81 (45.0%) |
| Positive | 71 (52.6%) | 28 (62.2%) | 99 (55.0%) |
| **CD4 T-cell count** |  |  |  |
| Mean (SD) | 317 (229) | 405 (278) | 341 (245) |
| **Household Food Insecurity Score** |  |  |  |
| Mean (SD) | 2.99 (5.05) | 4.16 (5.39) | 3.28 (5.15) |
| **Depression score** |  |  |  |
| Mean (SD) | 6.92 (4.94) | 10.10 (5.67) | 7.71 (5.30) |
| **Anxiety score** |  |  |  |
| Mean (SD) | 33.90 (8.88) | 37.10 (9.58) | 34.70 (9.14) |

*SD = Standard deviation*

**Figure 2.**
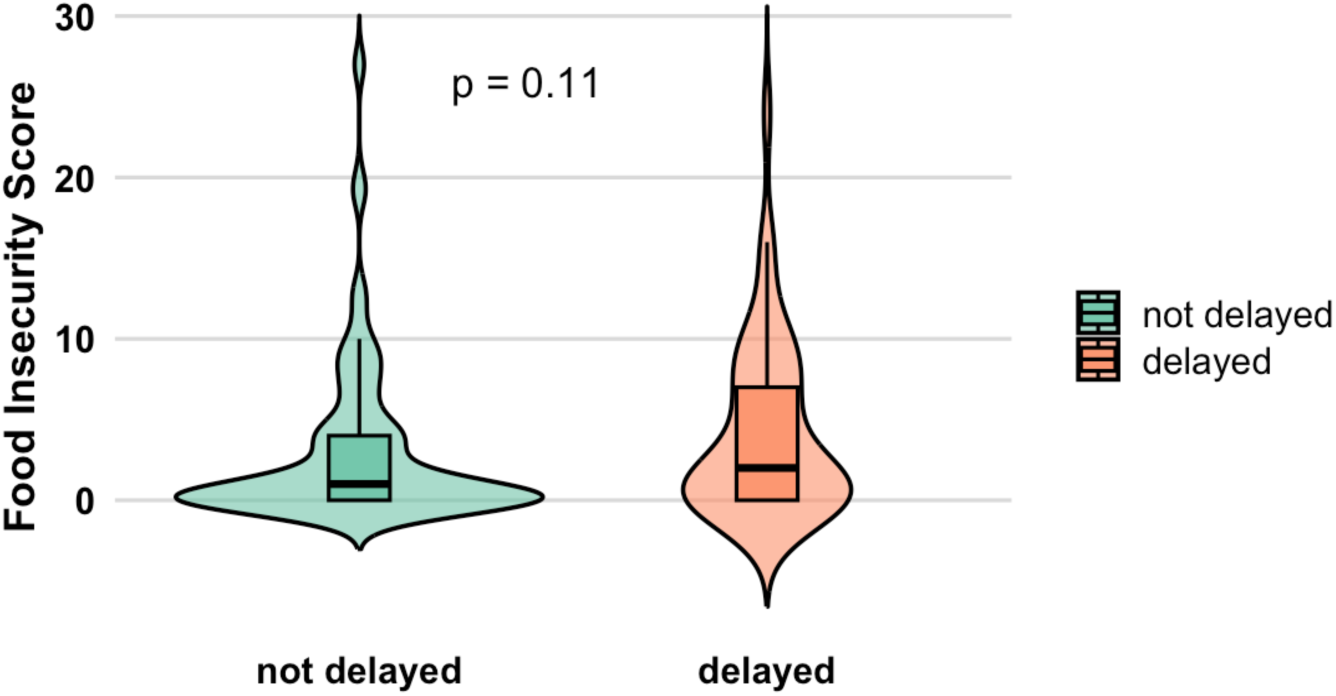
Distribution of Food Insecurity Scores by Treatment Initiation

**Figure 3.**
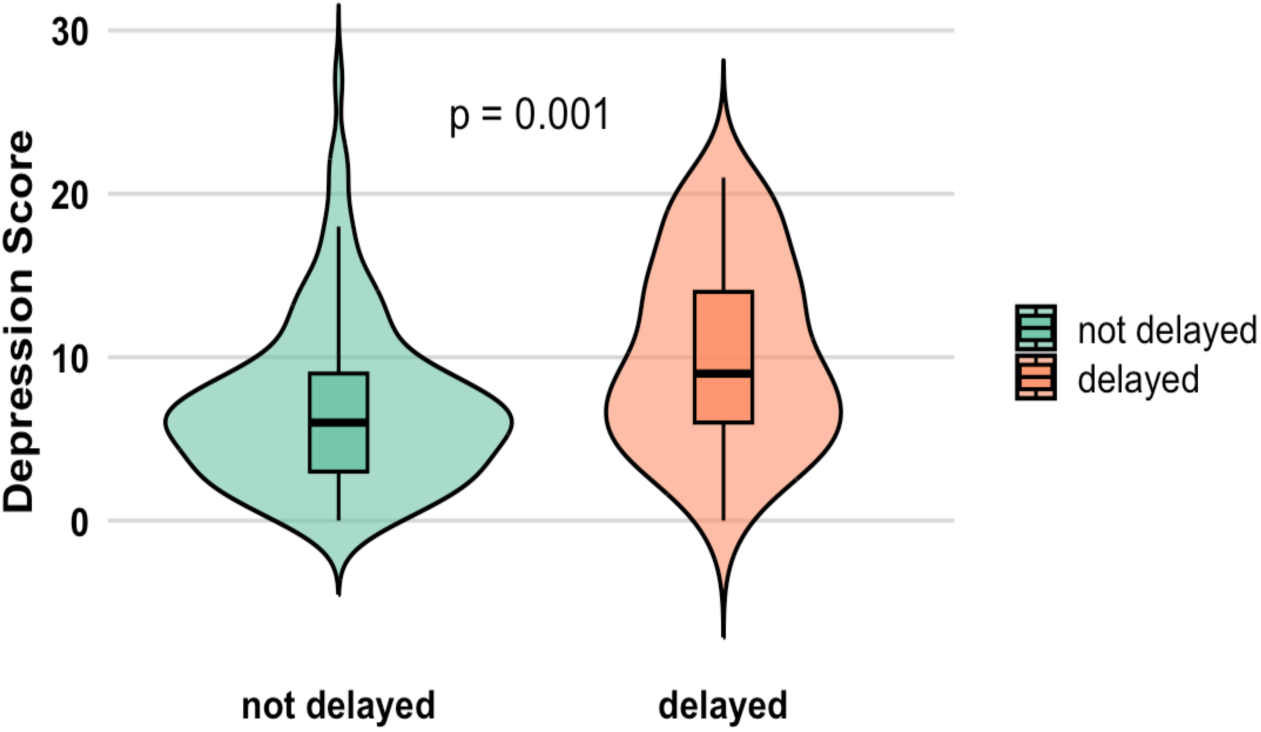
Distribution of Depression Scores by Treatment Initiation

**Figure 4.**
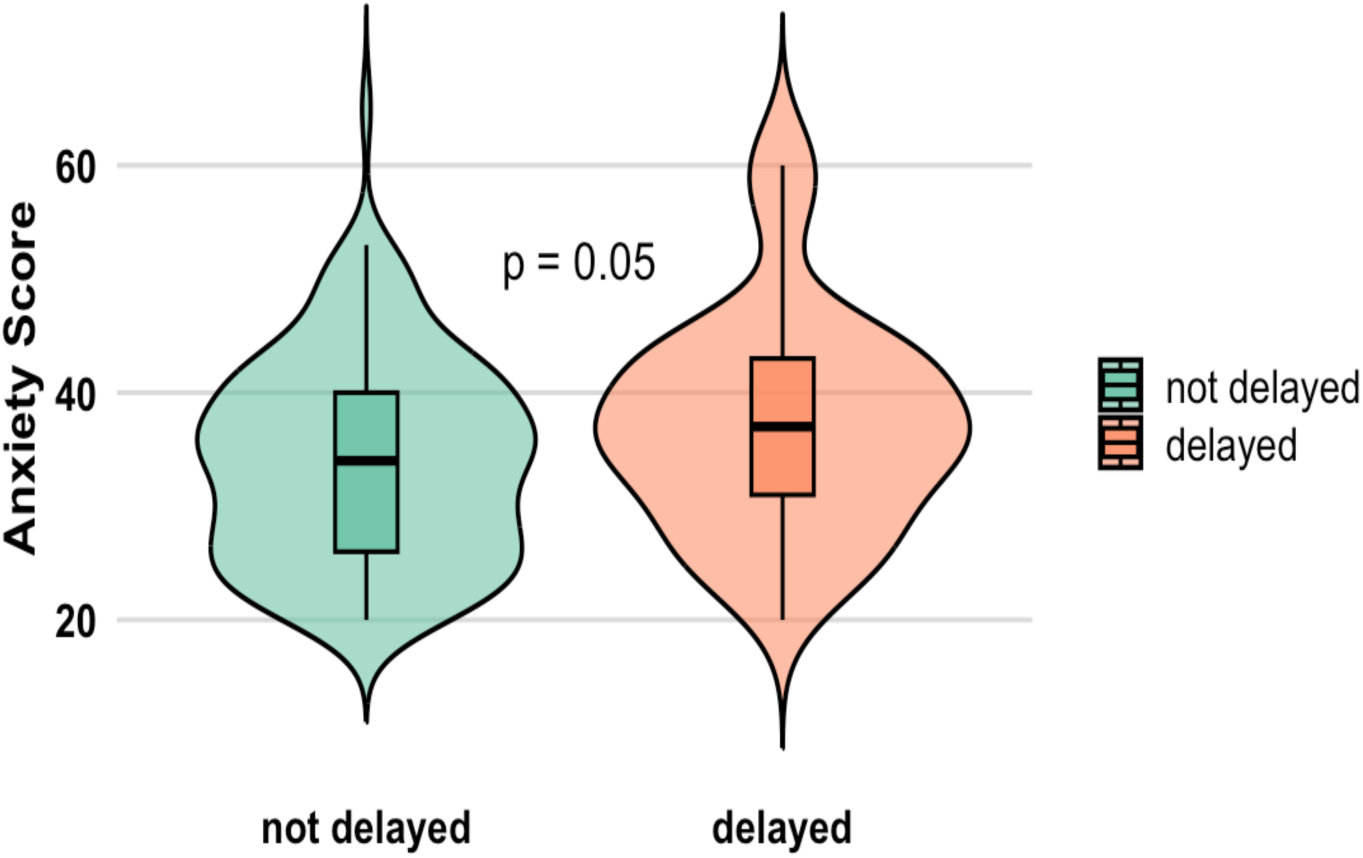
Distribution of Anxiety Scores by Treatment Initiation

There was insufficient evidence of association between food insecurity and delayed TB treatment initiation (OR = 1.04, 95% CI: 0.98 - 1.11, p = 0.20) from the standard regression model without considering mediation (**Table 2** which shows the results of the bivariate associations with delayed TB treatment). No other variable showed sufficient evidence of association with delayed TB treatment.

**Table 2:** Bivariate Associations with Delayed TB Treatment Initiation.

| Predictor Variable | OR | 95% CI | p-value |
| --- | --- | --- | --- |
| Household Food Insecurity Score | 1.04 | 0.98 - 1.11 | 0.20 |
| Age | 1.02 | 0.99 - 1.05 | 0.18 |
| Marital Status |  |  |  |
| Single | — | — |  |
| Married | 1.17 | 0.36 - 3.33 | 0.77 |
| **Sex** |  |  |  |
| Female | — | — |  |
| Male | 1.00 | 0.50 - 2.06 | >0.99 |
| **Educational status** |  |  |  |
| Less than secondary | — | — |  |
| Secondary or above | 0.91 | 0.45 - 1.81 | 0.79 |
| **Income status** |  |  |  |
| None | — | — |  |
| Yes | 1.03 | 0.52 - 2.12 | 0.93 |
| **HIV Status** |  |  |  |
| Negative | — | — |  |
| Positive | 1.48 | 0.75 - 3.01 | 0.26 |

CI = Confidence Interval, OR = Odds Ratio

**Table 3** presents the results for mediation analysis through depression. There was evidence of total natural indirect effect through depression (OR = 1.04, 95% CI: 1.01-1.08, p< 0.001) with 0.99 proportion mediated (p = 0.23). The first part of the indirect effect (**Figure 1**) showed that higher food insecurity was associated with higher depression scores (β = 0.37, 95% CI: 0.22 - 0.51, p < 0.001). The second part of the indirect effect also showed that higher depression score was associated with higher odds of experiencing delayed TB treatment (OR = 1.11, 95% CI: 1.05 - 1.19, p < 0.001). There was no evidence of a pure natural direct effect (OR = 1.00, 95% CI: 0.92 - 1.10, p = 0.97) or total effect (OR = 1.04, 95% CI: 0.97 - 1.13, p = 0.23).

**Table 3:** Mediation Effect of Depression between Food Insecurity and Delayed TB Treatment.

| Bivariate association between food insecurity and depression (path a) |  |  |  |  |
| --- | --- | --- | --- | --- |
|  | Beta | 95% CI | p-value |  |
| Household food insecurity score | 0.37 | 0.22 - 0.51 | <0.001 |  |
| Bivariate association between depression and delayed TB treatment initiation (path b) |  |  |  |  |
|  | OR | 95% CI | p-value |  |
| Depression | 1.11 | 1.05 - 1.19 | <0.001 |  |
| Mediation analysis results |  |  |  |  |
|  | estimate | Std. error | 95% CI | P-value |
| Pure Natural Direct effects | 1.00 | 0.04 | 0.92 - 1.10 | 0.97 |
| Total Natural Indirect effect | 1.04 | 0.02 | 1.01 - 1.08 | <0.001 |
| Total effects | 1.04 | 0.04 | 0.97 - 1.13 | 0.23 |
| Overall Proportion Mediated | 0.99 | - | - | 0.23* |

CI = Confidence Interval, OR = Odds Ratio, Std. error = Standard error
\* Because the total effect was close to the null value ( $OR \approx 1$ ), confidence interval and p-value estimates for proportion mediated are unstable.

The results for mediation analysis through anxiety are shown in **Table 4**. The results showed evidence of significant total natural indirect effects (OR = 1.03, 95% CI 1.01 - 1.06, p<0.001) through anxiety and significant total effects (OR = 1.08, 95% CI: 1.01 - 1.18, p=0.05). The overall proportion mediated was 0.38 (p=0.05). The first part of the indirect effect showed that higher food insecurity was associated with higher anxiety scores (β= 0.49, 95% CI: 0.24 - 0.74, p < 0.001). The second part of the indirect effect also showed that higher anxiety scores were associated with higher odds of experiencing delayed TB treatment (OR = 1.04, 95% CI: 1.00 - 1.08, p = 0.04). There was insufficient evidence of pure natural direct effect (OR = 1.05, 95% CI: 0.98 -1.14, p = 0.15). The results of the sensitivity analysis done by including HIV status in the final model were similar to those of the main analysis (**See Additional file 2)**.

**Table 4:** Mediation Effect of Anxiety between Food Insecurity and Delayed TB Treatment.

| Bivariate association between food insecurity and anxiety (path a) |  |  |  |  |
| --- | --- | --- | --- | --- |
|  | Beta | 95% CI | p-value |  |
| Household food insecurity score | 0.49 | 0.24 - 0.74 | <0.001 |  |
| Bivariate association between anxiety and delayed TB treatment initiation (path b) |  |  |  |  |
|  | OR | 95% CI | p-value |  |
| Anxiety | 1.04 | 1.00 - 1.08 | 0.04 |  |
| Mediation analysis results |  |  |  |  |
|  | estimate | Std. error | 95% CI | P-value |
| Pure natural direct effect | 1.05 | 0.04 | 0.98 - 1.14 | 0.15 |
| Total Natural indirect effect | 1.03 | 0.01 | 1.01 -1.06 | <0.001 |
| Total effects | 1.08 | 0.04 | 1.01 - 1.18 | 0.05 |
| Overall Proportion mediated | 0.38 | - | - | 0.05* |

CI = Confidence Interval, OR = Odds Ratio, Std. error = Standard error
\* Because the total effect was close to the null value ( $OR \approx 1$ ), confidence interval and $p$ -value estimates for proportion mediated are unstable.

## Discussion

This current study examined the mediating role of depression and anxiety for the relationship between food insecurity and delayed TB treatment. While standard regression analysis found insufficient evidence of an overall association between food insecurity and delayed TB treatment initiation, mediation analysis revealed significant indirect effects through depression and anxiety, suggesting that depression and anxiety may be possible pathways linking food insecurity to delayed TB treatment initiation. Sensitivity analysis also showed findings similar to the main analysis demonstrating that our findings were robust.

Our findings are conflict with an Ethiopian study which found no evidence of association between food insecurity and patient delay among people with TB attending public health facilities after adjusting for confounders (33). However, that study did not examine depression and anxiety as mediators. Our mediation results are broadly consistent with a qualitative study that linked food insecurity to delay in seeking TB care (34). Franke and colleagues identified food insecurity as one of the three major barriers to seeking TB care during the Covid pandemic(34).

Evidence from other studies suggest that mental health disorders may function as a link between food insecurity and poor health outcomes (35–37). A study among people with HIV-HCV coinfection in Canada found that depression partially mediated the relationship between severe food insecurity and risk of detectable HIV viral load (35). Similarly, Weiser et al. reported that mental health accounted for part of the relationship between food insecurity and worse HIV-related health outcomes (37). In that study, depressive symptoms accounted for 60.2% of the total effect of food insecurity on physical health status, supporting the role of mental health as a pathway linking food insecurity to adverse HIV outcomes (37).

Our results extend this literature by showing that depression and anxiety may also represent important pathways linking food insecurity and delayed TB treatment initiation. People experiencing unstable access to food in their household may experience chronic stress, which can increase the risk of mental health disorders including depression and anxiety (11,18,38,39). Symptoms of depression and anxiety including hopelessness, sleep disturbances, social isolation, excessive fear, and worry may (40) negatively affect motivation to seek TB care on time leading to delayed TB diagnosis and treatment (9,41). Kuznetsov et al. explained that people may be overcome with feelings of hopelessness, which can decrease the drive to seek TB care (9).

Food insecurity in Gaborone should be understood as a condition shaped by policies, structures and systems within Botswana and broader global geopolitics. In Gaborone, food insecurity has been linked to unemployment, unstable incomes, high energy cost, and a food system that relies heavily on large supermarkets and imported foods (40,42,43). Rapid urbanization in Gaborone, driven by the concentration of opportunities in urban settings and the relative underdevelopment of rural areas contributes to food insecurity (40,42). Increased migration into the urban Gaborone, increases pressure on job opportunities, leading to high unemployment and reliance on informal sources of income (40,42,44). Household energy costs also represent an important structural barrier that negatively impacts food security among urban households. Low-income households experience difficulties in affording electricity and fuel needed for storage and preparation of food considered nutritious but may require longer cooking times (40). These conditions may reduce household purchasing power and make it difficult for people to maintain consistent access to food, especially during periods of illness where people may not be able to work (40,45). Additionally, limited agricultural production in urban Gaborone contributes to dependance on imported foods and large supermarkets. Policies to assist and strengthen local food production remains limited or non-existent, increasing reliance on external food sources (42,43). Policies have placed little attention to urban food security (40,43). For instance, people with TB may be offered food packages as part of their treatment to help address their nutritional needs. However, such interventions may be underfunded, insufficient to support the household, and ineffective in addressing the needs of vulnerable urban households (15). Among people living with TB and HIV, these structures compound existing vulnerabilities, further limiting the quantity, quality, availability, and access to nutritious food on a consistent basis.

The findings of the current study show the need for further research to better understand the pathways linking food insecurity to delayed TB treatment. Studies with larger samples and longitudinal designs are needed to confirm temporality of depression and anxiety and their mediating roles with food insecurity and delayed TB treatment. The study results also suggest that interventions aimed at improving TB treatment should integrate mental health services with social and financial support considering that the need for social protection against food insecurity and other economic stressors is likely to increase substantially given the recent decline in Botswana’s economy driven by reduction in the global diamond market (46). Population level interventions aimed at addressing household food insecurity such as cash transfers and food assistance programs may have benefits beyond nutrition alone because it may also reduce depression and anxiety that could interfere with seeking TB treatment and adherence. Our study builds on mounting evidence that shows that improved TB care cannot be fully achieved without addressing social and structural determinants of health (47,48).

## Strengths and limitations

This study focused on people living with TB in a marginalized population. To the best of our knowledge, no previous study has examined the mediating role of depression and anxiety for the relationship between food insecurity and delayed TB treatment initiation. This current study also has several limitations. First, the sample size was relatively small, with only 45 delayed TB treatment initiation events, which limited statistical power and our ability to adjust for a larger number of confounders. Second, the cross-sectional design limits the ability to establish temporality and causality, which are important in mediation analysis. Our mediation analysis also assumed no unmeasured confounding, which may not represent actual conditions. Finally, delayed TB treatment was defined using total delay time only. Other forms of TB treatment delay, including patient delay and health system delay, were not assessed in this study.

## Conclusion

This study found an association between food insecurity and delayed TB treatment initiation mediated by indirect effects through depression and anxiety. This indirect effect suggests that mental health disorders may represent important pathways that link food insecurity to delayed TB treatment initiation. These findings add to the limited literature in this area and highlights the importance of considering both socioeconomic hardships and psychological factors in addressing delayed TB treatment.

## Declarations

## Data Availability

All data produced in the present study are available upon reasonable request to the corresponding author.

## Acknowledgement

We express our gratitude to all research staff and participants who were involved in this study.

## Ethics approval and consent to participate

Ethical approval for the study was obtained from the Institutional Review Boards at the Botswana Ministry of Health and Wellness and University of California Irvine. This study was conducted according to the guidelines of the Helsinki Declarations. Clinical trial number: Not applicable.

## Authors’ contributions

E.S. contributed to study design, data analysis and led the manuscript writing. K.M. contributed to data collection. C.M. contributed to study supervision. S.S.S. contributed to study design, supervised the analysis and manuscript writing. All authors read and approved final manuscript.

## Availability of data and materials

Dataset is available upon reasonable request from the corresponding author.

## Consent for publication

Not applicable

## List of abbreviations

CD-4: Cluster of Differentiation 4
CI: Confidence Interval
HCV: Hepatitis C virus
HFIAS: Household food insecurity Access Scale
HIV: Human immunodeficiency virus
PHQ-9: Patient Health Questionnaire 9
OR: Odds Ratio
SD: Standard deviation
Std. error: Standard error
TB: Tuberculosis
USAID: United States Agency for International Development
WHO: World Health Organization

