## Additional file 1 for "Mediating Role of Depression and Anxiety in the Association Between Food Insecurity and Delayed TB Treatment in Botswana: A Cross-Sectional Study"

AGReMA
A Guideline for Reporting Mediation Analyses

| Section/Topic | Item Number | Item Description | Reported on page No |
|---|---|---|---|
| **Title and abstract** | | | |
| Title | 1 | Identify that the study uses mediation analysis | 1 |
| Abstract | 2 | Provide a structured summary of the objectives, methods, results, and conclusions specific to mediation analyses | 2,3 |
| **Introduction** | | | |
| Background and rationale | 3 | Describe the study background and theoretical rationale for investigating the mechanisms of interest. Include supporting evidence or theoretical rationale for why the intervention or exposure might have a causal relationship with the proposed mediators. Include supporting evidence or theoretical rationale for why the mediators might have a causal relationship with the outcomes | 4,5 |
| Objectives | 4 | State the objectives of the study specific to the mechanisms of interest. The objectives should specify whether the study aims to test or estimate the mechanistic effects | 5 |
| **Methods** | | | |
| Study registration | 5 | If applicable, provide references to any protocols or study registrations specific to the mediation analysis, and highlight any deviations from the planned protocol | Not applicable |
| Study design and source of data | 6 | Specify the design of the original study that was used in mediation analyses and where the details can be accessed, supported by a reference. If applicable, describe study design features that are relevant to mediation analyses | 5 |
| Participants | 7 | Describe the target population, eligibility criteria specific to mediation analyses, study locations, and study dates (start of participant enrolment and end of follow-up) | 5, 6 |
| Sample Size | 8 | State whether a sample size calculation was conducted for mediation analyses. If so, explain how it was calculated | Not calculated. Data had already been collected before we did analysis. |
| Effects of interest | 9 | Specify the effects of interest | 9 |
| Assumed causal model | 10 | Include a graphic representation of the assumed causal model including the exposure, mediator, outcome, and possible confounders | 8 |
| Causal assumptions | 11 | | 9,10 |
| Measurement | 12 | Clearly describe the interventions or exposures, mediators, outcomes, confounders, and moderators that were used in the analyses. Specify how and when they were measured, the measurement properties, and whether blinded assessment was used | 6,7,8 |


(cc) BY-NC-ND

| | | | |
|---|---|---|---|
| Measurement levels | 13 | If relevant, describe the levels at which the exposure, mediator, and outcome were measured | Not applicable |
| Statistical methods | 14 | Describe the statistical methods used to estimate the causal relationships of interest. This description should specify analytical strategies used to reduce confounding, model building procedures, justification for the inclusion or exclusion of possible interaction terms, modelling assumptions, and methods used to handle missing data. Provide a reference to the statistical software and package used | 7,8,9,10 |
| Sensitivity analyses | 15 | Describe any sensitivity analyses that were used to explore causal or statistical assumptions and the influence of missing data | 9 |
| Ethical approval | 16 | Name the institutional research board or ethics committee that approved the study. Provide a description of participant informed consent or ethics committee waiver of informed consent | 6 |
| **Results** | | | |
| Participants | 17 | Describe baseline characteristics of participants included in mediation analyses. Report the total sample size and number of participants lost during follow-up or with missing data | 10 |
| Outcomes and estimates | 18 | Report point estimates and uncertainty estimates for the exposure-mediator and mediator-outcome relationships. If inference concerning the causal relationship of interest is considered feasible given the causal assumptions, report the point estimate and uncertainty estimate | 14,15,16 |
| Sensitivity parameters | 19 | Report the results from any sensitivity analyses used to assess robustness of the causal or statistical assumptions, and the influence of missing data | 15 |
| **Discussion** | | | |
| Limitations | 20 | Discuss the limitations of the study including potential sources of bias | 19 |
| Interpretation | 21 | Interpret the estimated effects considering the study's magnitude and uncertainty, plausibility of the causal assumptions, limitations, generalizability of the findings, and results from relevant studies | 16,17,18,19 |
| Implications | 22 | Discuss the implications of the overall results for clinical practice, policy, and science | 20 |
| **Other information** | | | |
| Funding and role of sponsor | 23 | List all sources of funding or sponsorship for the mediation analysis and the role of the funders/sponsors in the conduct of the study, writing of the manuscript, and decision to submit for publication. | 21 |
| Conflicts of interest and financial disclosures | 24 | State any conflicts of interest and financial disclosures for all authors | 21 |
| Data and code | 25 | Authors are encouraged to provide a statement for sharing data and code for the mediation analysis | 21 |

*From:* Lee H, Cashin AG, Lamb SE, Hopewell S, Vansteelandt S, VanderWeele TJ, et al. A Guideline for Reporting Mediation Analyses of Randomized Trials and Observational Studies. The AGReMA Statement. JAMA. 2021;326(11):1045–1056. doi:10.1001/jama.2021.14075


(cc) BY-NC-ND

AGReMA
A Guideline for Reporting Mediation Analyses

AGReMA is designed for articles that report mediation analyses of randomized trials or observational studies.
For more information, visit: agrema-statement.org


(cc) BY-NC-ND
