## Additional file 2 for "Mediating Role of Depression and Anxiety in the Association Between Food Insecurity and Delayed TB Treatment in Botswana: A Cross-Sectional Study"

**Figure 6**

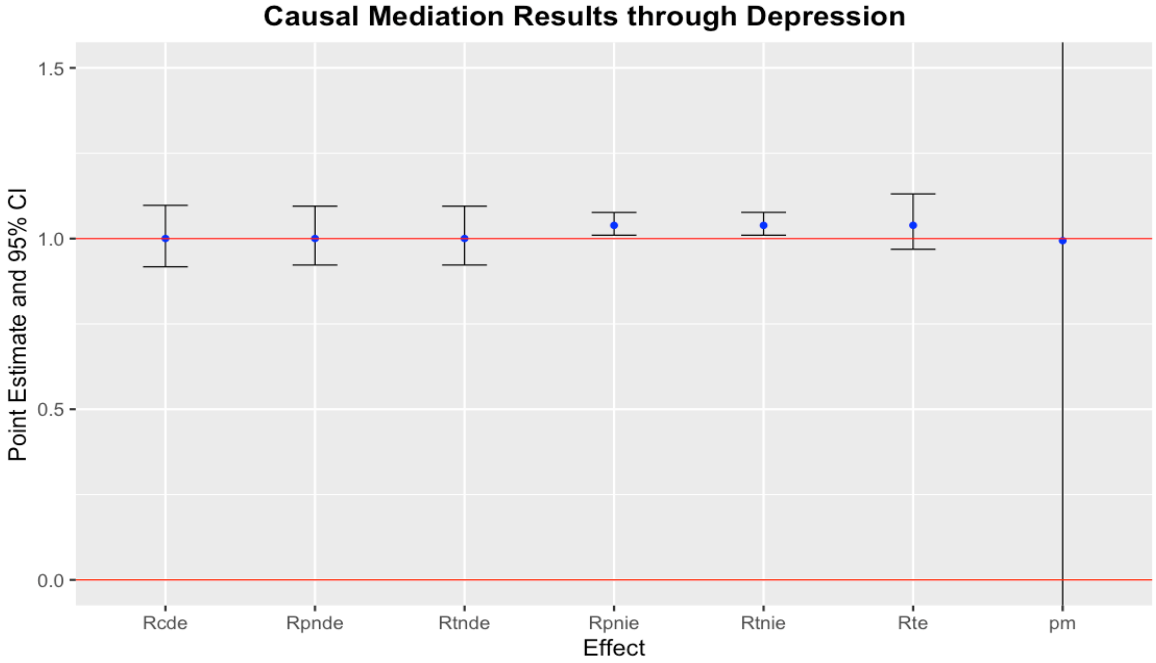


*Rcde: controlled direct effect odds ratio; Rpnde: pure natural direct effect odds ratio; Rtnde: total natural direct effect odds ratio; Rpnie: pure natural indirect effect odds ratio; Rtnie: total natural indirect effect odds ratio; Rte: total effect odds ratio; pm: overall proportion mediated*

**Figure 6** shows the results of mediation analysis through depression for the association between food insecurity and delayed TB treatment. Confidence interval for Proportion mediated (pm) should be interpreted with caution because total effects odds ratio is close to the null value (OR =1) hence making confidence interval for pm unstable.

**Figure 7**

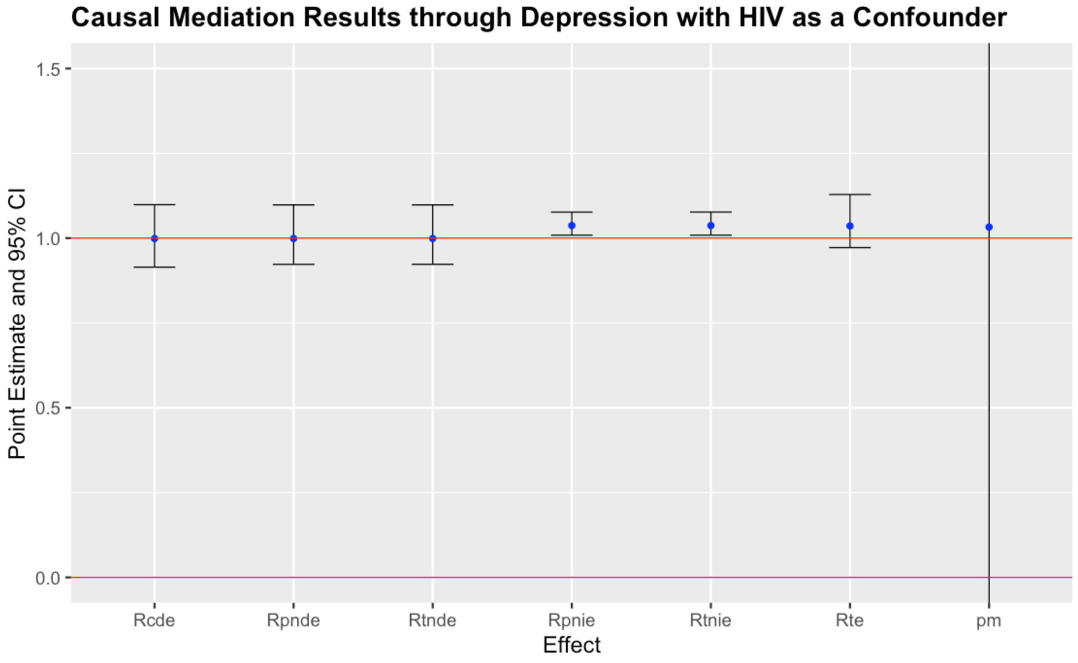


*Rcde: controlled direct effect odds ratio; Rpnde: pure natural direct effect odds ratio; Rtnde: total natural direct effect odds ratio; Rpnie: pure natural indirect effect odds ratio; Rtnie: total natural indirect effect odds ratio; Rte: total effect odds ratio; pm: overall proportion mediated*

Figure 7 shows the results of sensitivity analysis when HIV status was included as confounder in the mediation model for depression. Confidence interval for Proportion mediated (pm) should be interpreted with caution because total effects odds ratio is close to the null value ( OR =1) hence making confidence interval for pm unstable.

**Figure 8**

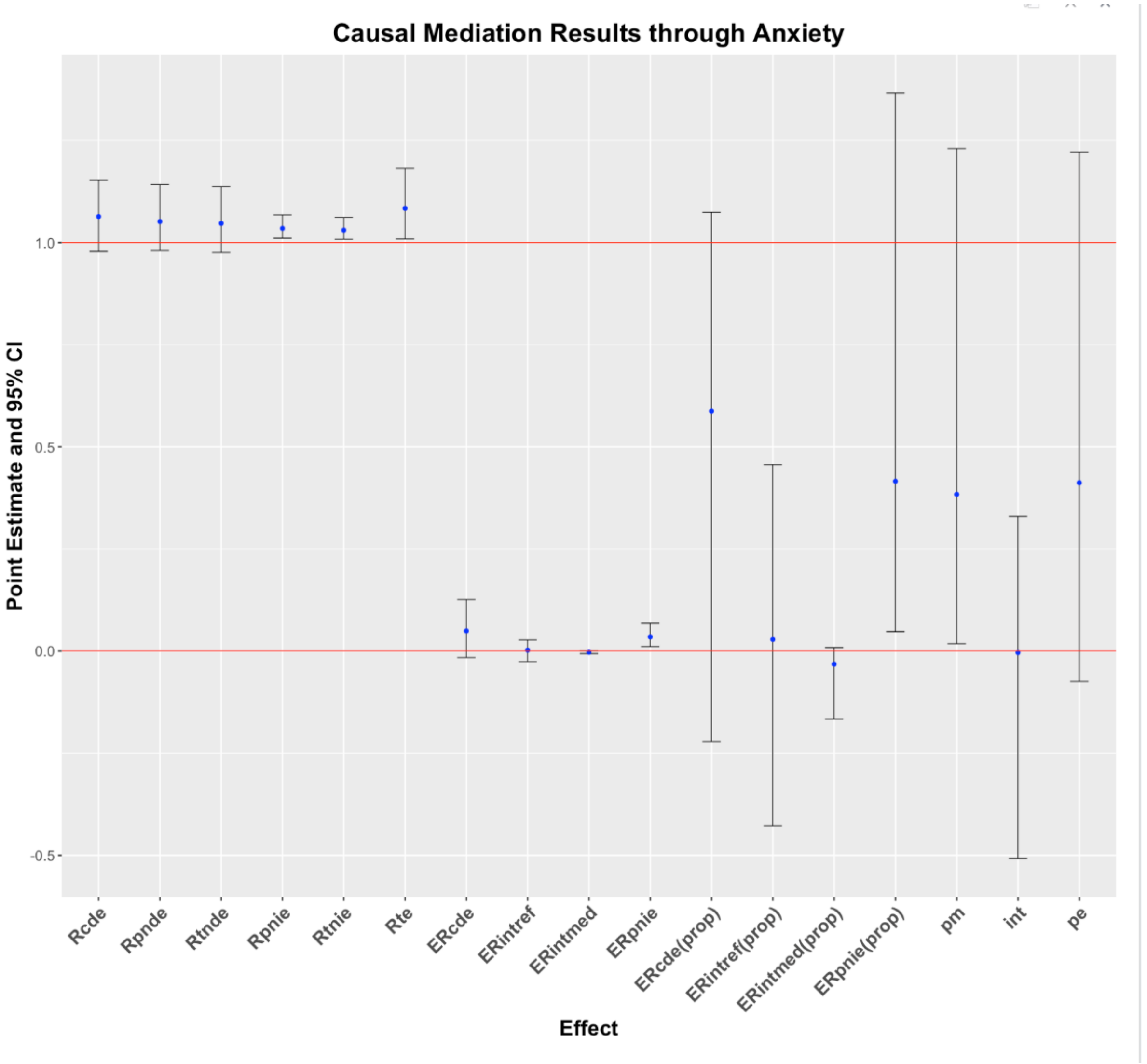


*Rcde: controlled direct effect odds ratio; Rpnde: pure natural direct effect odds ratio; Rtnde: total natural direct effect odds ratio; Rpnie: pure natural indirect effect odds ratio; Rtnie: total natural indirect effect odds ratio; Rte: total effect odds ratio; ERcde: excess relative risk due to controlled direct effect; ERintref: excess relative risk due to reference interaction; ERintmed: excess relative risk due to mediated interaction; ERpnie: excess relative risk due to pure natural indirect effect; ERcde(prop): proportion ERcde; ERintref(prop): proportion ERintref; ERintmed(prop): proportion ERintmed; ERpnie(prop): proportion ERpnie; pm: overall proportion mediated; int: overall proportion attributable to interaction; pe: overall proportion eliminated*

**Figure 8** shows the results of mediation analysis through anxiety for the association between food insecurity and delayed TB treatment. Confidence interval for Proportion mediated (pm) should be interpreted with caution because total effects odds ratio is close to the null value (OR =1) hence making confidence interval for pm unstable.

**Figure 9**

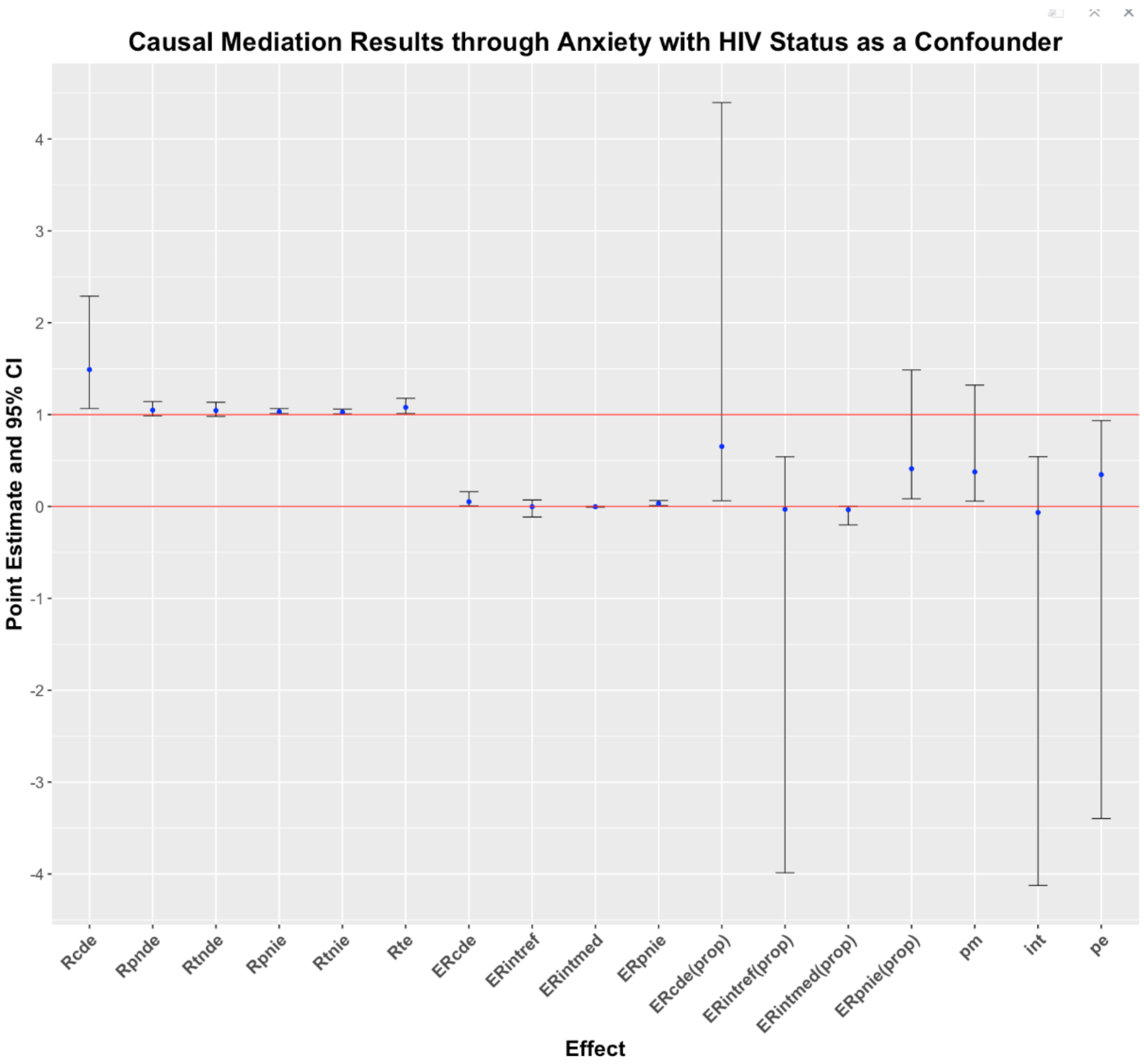


*Rcde: controlled direct effect odds ratio; Rpnde: pure natural direct effect odds ratio; Rtnde: total natural direct effect odds ratio; Rpnie: pure natural indirect effect odds ratio; Rtnie: total natural indirect effect odds ratio; Rte: total effect odds ratio; ERcde: excess relative risk due to controlled direct effect; ERintref: excess relative risk due to reference interaction; ERintmed: excess relative risk due to mediated interaction; ERpnie: excess relative risk due to pure natural indirect effect; ERcde(prop): proportion ERcde; ERintref(prop): proportion ERintref; ERintmed(prop): proportion ERintmed; ERpnie(prop): proportion ERpnie; pm: overall proportion mediated; int: overall proportion attributable to interaction; pe: overall proportion eliminated*

Figure 9 shows the results of sensitivity analysis when HIV status was included as confounder in the mediation analysis model for anxiety. Confidence interval for Proportion mediated (pm) should be interpreted with caution because total effects odds ratio is close to the null value (OR =1) hence making confidence interval for pm unstable.
